# Feasibility and reliability of online vs in-person cognitive testing in healthy older people

**DOI:** 10.1101/2023.07.05.23292229

**Authors:** Sol Morrissey, Rachel Gillings, Michael Hornberger

## Abstract

**Background:** Early evidence in using online cognitive assessments show that they potentially offer a feasible and resource efficient alternative to in-person clinical assessments in evaluating cognitive performance, yet there is currently little understanding about how these assessments relate to traditional, in-person cognitive tests.

**Objectives:** We assess the feasibility and reliability of NeurOn, a novel online cognitive battery, measuring processing speed, executive functioning, spatial working memory, episodic memory, attentional control, visuospatial functioning and spatial orientation.

**Design:** 32 participants (mean age: 70.19) completed two testing sessions, unsupervised online and in-person, one-week apart. Participants were randomised in the order of testing appointments. For both sessions, participants completed questionnaires prior to a cognitive assessment. Test-retest reliability and concurrent validity of the online cognitive battery was assessed by comparing performance in repeated tasks across testing sessions as well as with traditional in-person cognitive tests.

**Results:** Global cognition in the NeurOn battery moderately validated against MoCA performance. The battery demonstrated moderate test-retest reliability as performance across repeated tasks did not show a significant difference. Concurrent validity was found only between the online and paper versions of the Trail Making Test -A, as well as global cognitive performance between online and in-person testing sessions.

**Conclusions:** The NeurOn cognitive battery provides a promising tool for measuring cognitive performance online both longitudinally and across short retesting intervals within healthy older adults. When considering cost-effectiveness, flexible administration, and improved accessibility for wider populations, online cognitive assessments have the potential to improve future screening for neurodegenerative diseases.

## Introduction

Cognitive functioning is vital for everyday behaviour. It is well established that within ageing, many aspects of cognition typically decline (1,2). It is also increasingly evident that subtle cognitive changes appear long before clinical manifestations of neurodegenerative diseases such as dementia become apparent (3). Therefore, it is vital to understand whether these cognitive changes are indicative of neurodegenerative disease symptomatology or are typical of healthy ageing. Earlier diagnosis of cognitive impairment is invaluable for patients and caregivers as it can inform management of patient wellbeing and provide targeted measures for lifestyle modifications to potentially reverse cognitive decline (4,5).

Neuropsychological testing is therefore required to measure changes in cognitive functioning (6,7). Nonetheless, routine cognitive assessments in healthy ageing are rarely conducted and typically rely upon quick to administer paper-based tests (8). The current gold-standard tests for assessing cognitive impairment, such as the Mini-Mental State Examination (MMSE), were developed to screen for dementia, but are less sensitive in identifying milder cognitive impairment (9,10). Furthermore, clinic assessments involving paper-based tests are limited as they are prone to practice effects (11) and cognitive changes may be masked by fluctuations in cognitive performance or differences in cognitive reserve (12).

In recent years, exciting developments in online cognitive testing have increased its usage in both research and clinical environments (13). Importantly, online assessments can be performed remotely to improve accessibility and frequency of online cognitive testing, enabling identification of more subtle changes in cognitive decline (14,15). Digital assessments also provide enhanced precision in data measurement, standardised presentation, pseudorandomisation to reduce practice effects, and greater cost-efficiency (13,16). To date, the majority of computerised testing has focussed upon processing speed and attention tasks, with many demonstrating promising results (17–19). Similarly, a recent systematic review found that early evidence of computerised cognitive testing shows potential clinical utility in diagnosing neurocognitive disorders, yet there has been little validation work in cognitive batteries thus far – which is necessary to establish whether they are feasible for clinical applications (20).

Comprehensive neuropsychological test batteries testing a variety of domains are required to detect early cognitive changes that may manifest in older age. Normative data for healthy older adults is necessary to enable for interpreting cognitive performance in context of sociodemographic factors, so that at-risk populations can be accurately identified. Our group recently developed NeurOn, a novel cognitive battery, as part of the DECISION study (Morrissey et al., unpublished). NeurOn is a comprehensive cognitive battery testing a variety of cognitive domains and is novel in that it also assesses spatial orientation ability – of which previous work from our group has shown to be a key signature for preclinical dementia (21).

In the present study, we aim to evaluate the psychometric properties of the NeurOn battery by measuring the reliability and validity in both supervised in-person and unsupervised online settings against established traditional neuropsychological assessments. It is hypothesised that online cognitive tasks will demonstrate test-retest reliability over a one-week period; online/remote cognitive tasks will demonstrate concurrent validity with in-person/traditional cognitive task equivalents; and that cognitive performance in the neuropsychological battery will validate against established clinical tests in measuring cognitive performance.

## Methods

### Recruitment

33 older adults (65+) were recruited to take part in an experimental study. All participants were pre-screened to assess whether they were cognitively and physically healthy; had any history of psychiatric or neurological disease; history of substance abuse disorder; drive once per week or more; and whether they had previously taken part in a study using the online cognitive platform. Participants underwent testing between October 2022 and March 2023. Signed informed consent was obtained from each participant and data was attributed anonymously. Ethical approval for the study was provided by the Faculty of Medicine and Health Sciences Research Ethics Committee at the University of East Anglia (FMH2019/20-134).

### Procedure

Screening was carried out via online video call (32) and telephone (1) by the study team prior to baseline cognitive assessment. One participant was excluded from the study as they only completed one testing session due to illness. Participants were randomised to the order in which they completed testing sessions. Prior to the baseline appointment, participants were asked with which device they would most comfortably complete the remote assessment appointment (desktop, laptop, tablet) and the device was matched for the in-person testing appointment. Both testing sessions started with completion of questionnaires pertaining to demographics, subjective cognition, and driving history. Each participant completed the follow-up testing session one week from the baseline testing session at the same time of day.

### Development and description of the online cognitive testing platform

Questionnaires and cognitive tasks were hosted on NeurOn – an online platform. The novel cognitive battery was developed by a professional programmer alongside the project team. Online neuropsychological tests were based on a combination of established, traditional neuropsychological tests and established novel tasks (VST) and was developed for unsupervised assessment. Tests were designed to be completed in unmonitored conditions. Tasks were accompanied with written instructions and video tutorials with a voice-over (except for Go-No/Go test) prior to test completion to promote multimodal learning. After receiving instructions, practice sessions for each task followed to ensure participants were prepared for the real test. Participants were encouraged to complete the main test battery in one session without breaks but were encouraged to take a break prior to the Virtual Supermarket Task due it having a significantly longer duration and greater task difficulty. If the cognitive test battery was interrupted (i.e. by participants taking a break/ internet disconnection), upon logging back into the battery, participants resumed the task from their current progress. All tasks were pseudorandomised to enable for repeated testing. All participant input was saved on a protected server throughout each test element.

### Online cognitive tasks

The NeurOn battery consisted of a variety of digitalised tasks measuring cognition across various domains. A Reaction Time task, whereby participants responded as quickly as possible to a repeating on-screen stimulus, measured visuomotor speed. Trail-Making Test - A, involving the connecting of 25 numerically arranged points in ascending order as quickly as possible, measured processing speed. Trail-Making Test - B, involving the connecting of 25 points of alternating numbers and letters in ascending order as quickly as possible, measured executive functioning. Picture Recognition involved a stimulus encoding phase of everyday objects appearing consecutively in varying screen locations prior to a delayed testing phase, where participants were asked whether an appearing stimuli appeared previously (measuring recognition memory), and if so at which screen position (measuring source memory). A Spatial Span – Backwards task measured spatial working memory, whereby participants are presented with an array of boxes that light up in an ascending sequential order per trial (2–9) and then must relay the previous sequence in reverse order. The Go-No/Go task measured attentional control by asking participants to respond to a specific stimuli (Go) and inhibit responses to other stimuli (No-go); Fragmented Letters assessed visuospatial functioning by asking participants to identify a singular letter from the alphabet which is fragmented through a visual mask. Finally, participants completed the Virtual Supermarket Task, previously described in detail (22), which measured for allocentric and egocentric orientation by asking participants to orientate a trolley in a virtual supermarket according to a previously presented video clip.

### Remote cognitive testing

Participants completed the remote cognitive testing session from their own home. Initially, participants completed demographics and novel subjective cognition questionnaires (Spatial Memory & Driving, Orienteering, and Navigation). The online cognitive test battery consisted of Reaction Time, Trail Making Test - A, Trail Making Test - B, Picture Recognition, Spatial Span Backwards, Go-No-Go test, Fragmented Letters, and Virtual Supermarket Test.

### In-person cognitive testing

The in-person cognitive testing session took place in a quiet testing facility and involved a combination of traditional neuropsychological tests which require face-to-face assessment with our novel online tasks. Participants initially completed established questionnaires measuring subjective cognition (Cognitive Change Index (CCI) (23) and Santa Barbara Sense of Direction (SBSOD) (24)). Participants then completed the Montreal Cognitive Assessment (MoCA) (25), Reaction Time (Online), paper versions of the Trail-Making Test A & B (26), Rey Osterrieth Complex Figure Test (ROCF) – delayed recall (27), Corsi Block Tapping Test (28), Go-No/Go (Online), a paper version of the Fragmented Letters test (29), and lastly the Virtual Supermarket Task (Online).

### Statistical analyses Neuropsychological test measures

Extreme outlier cases were identified via boxplots. For the online session, outliers were removed for Reaction Time (1), Trail Making Test - A (1), Recognition Memory (1). For the in-person testing session, outliers were removed for the CCI (1), Reaction Time (2), Trail Making Test - A (1), Trail Making Test - B (1), ROCF recall (2), and Go-No/Go (1). Two participants did not complete the Virtual Supermarket Task in either test session due to either a technical error or finding the task too difficult. To create an episodic memory measure for the online cognitive battery, an average score was found between recognition and source memory percentages for each participant. A Bonferroni adjusted significance level of 0.00625 (0.05/8) was used to assess statistical significance in correlations between the CCI and online cognitive assessments. Raw cognitive test scores were standardised for regression analysis, except for episodic memory which was converted into a proportion as this score measured for accuracy in percentage.

### Concurrent validity (remote vs in-person testing)

Concurrent validity was measured by Spearman or Pearson correlations (depending on variable distribution) to assess the consistency between remote/online and in-person/traditional neuropsychological tests. A correlation threshold of ≥0.40 was used to establish acceptable concurrent validity (30). The online Trail Making Tests were compared to the paper Trail-Making Tests; the Spatial Span-Backwards task was compared with the Corsi Block Tapping test; the Picture Recognition task was compared with the ROCF-delayed recall task; and Fragmented Letters was compared with the paper Fragmented Letters task.

### Test-retest reliability

To examine test-retest reliability of the repeated online cognitive tasks from baseline to retest sessions, two complimentary approaches were conducted:

1. Two-way mixed effects intraclass correlation coefficients (ICCs) with measures of absolute agreement (95% CI) according to McGraw & Wong (31).
2. Paired samples *t* tests assessed performance differences. A significant (*p* <.05) improvement over time was used as a threshold to indicate practice effects.

### Global cognition

To establish a global cognition score for each testing session, Z-scores for each neuropsychological measure within each testing session were averaged to create a composite score. Z-scores were reversed to ensure consistent directionality within each task.

## Results

### Demographics

32 participants (15 female) were recruited for the study. The average age of participants was 70.19 ± 4.84, and the average years spent in education was 15.93 ± 3.94. In devices used, 41% of participants used desktops, 41% used laptops, and 18% used tablet devices to complete the study. On average, the online testing session took 58 minutes and 50 seconds whilst the in-person testing session took 66 minutes and 50 seconds. No significant differences were found between age, education or time taken to complete online and in-person testing batteries between males and females.

### Concurrent validity (remote vs. in-person testing)

To determine how online cognitive tests validated against traditional cognitive tests, concurrent validity was measured for online tasks with traditional cognitive test equivalents. Only Trail Making Test - B met the acceptable correlation threshold value to demonstrate acceptable concurrent validity between tasks, *r*(28) = 0.615, *p* <.001. Low correlations were established for Trail Making Test - A (0.25), Spatial Working Memory (0.27), and Episodic Memory (0.24). A ceiling effect was observed for the Fragmented Letters task in both paper and online versions across both testing sessions (see Table 1).

**Table 1.** Concurrent validity between online tasks and traditional neuropsychological tests.

| Test | Online (M (SD)) | Traditional (M(SD)) | Spearman's $\rho$ (S)/<br>Pearson's $r$ |
| --- | --- | --- | --- |
| Trail-Making Test -A | 32.13 (77.87) | 35.67 (12.67) | 0.25 |
| Trail-Making Test -B | 50.28 (19.16) | 69.19 (19.47) | 0.61*** |
| Spatial Working Memory | 5.44 (0.98) | 5.56 (1.24) | 0.27 |
| Episodic Memory | 90.75 (8.19) | 20.88 (4.27) | 0.24 (S) |
| Fragmented Letters | 100 (0) | 100 (0) | - |
| Global cognition | 0.22 (0.38) | 27.13 (2.03) | 0.60** |

*Note.*
\* $p < .05$ , \*\* $p < .01$ , \*\*\* $p < .001$
† Traditional tests of online tasks: Trail Making Tests: Trail Making Tests (paper versions), Spatial-Working Memory: Corsi Block Tapping Test; Episodic Memory: ROCF-delayed recall; Fragmented Letters: Fragmented Letters (paper); Global cognition: MoCA score.
†† Global cognition was calculated by averaging reversed Z scores for main neuropsychological tasks in online and in-person test settings.
††† Spearman's $\rho$ was used for Episodic Memory correlation as the online episodic memory score showed a non-normal distribution.

### Test-retest reliability and practice effects

Across all four repeated tasks, intraclass correlation coefficients demonstrated moderate test-retest reliability (0.50-0.80). Correlation coefficients ranged from 0.51 (Go/No-Go) to 0.75 (Egocentric Orientation). No practice effects were found for Reaction Time (online: M = 344.62 ± 69.02; in-person: M = 348.39 ± 72.89); Go/No-Go (online: M = 1.22 ± 1.64, in-person: M = 1.42 ± 1.36); Allocentric Orientation (online: M = 3.11 ± 1.57, in-person: M = 3.11 ± 1.62); or Egocentric Orientation (online: M = 51.31 ± 34.16, in-person: M = 56.52 ± (36.37), as all *t* test values were insignificant (*p* >.05) (see Table 2).

**Table 2.** Test-retest reliability of cognitive tasks between online and in-person testing sessions.

|  |  |  |  |  |  |
| --- | --- | --- | --- | --- | --- |
| **Test** | **Online (M (SD)** | **In-Person (M(SD)** | **Practice effects**<br>**(*t* (df))** | **ICC** | **Spearman's $\rho$ (S)/**<br>**Pearson's *r*** |
| Reaction time | 344.62 (69.02) | 348.39 (72.89) | -0.50 (29) | 0.55*** | 0.54** |
| Go/No-go | 1.22 (1.64) | 1.42 (1.36) | -0.60 (30) | 0.51** | 0.50** (S) |
| Allocentric<br>orientation | 3.11 (1.57) | 3.11 (1.62) | 0.11 (26) | 0.64*** | 0.63*** |
| Egocentric<br>orientation | 51.31 (34.16) | 56.52 (36.37) | -0.68 (28) | 0.75*** | 0.74*** |
| *Note.*<br>*$p < .05$, **$p < .01$, ***$p < .001$<br>† Spearman's $\rho$ was used for Go/No-go correlation as the online episodic memory score showed a non-normal distribution. |  |  |  |  |  |

### Association with established cognitive assessments

To determine how the online cognitive testing battery is associated with established cognitive assessments, correlation analysis was carried out between individual cognitive tests and total CCI score. Spearman rank correlation analysis found that higher CCI score was positively associated with worse egocentric orientation performance, *r*(27) = −.453, *p* = .014, however this was not statistically significant after Bonferroni correction. No other cognitive assessments were found to correlate with the CCI (see Table 3).

**Table 3.**
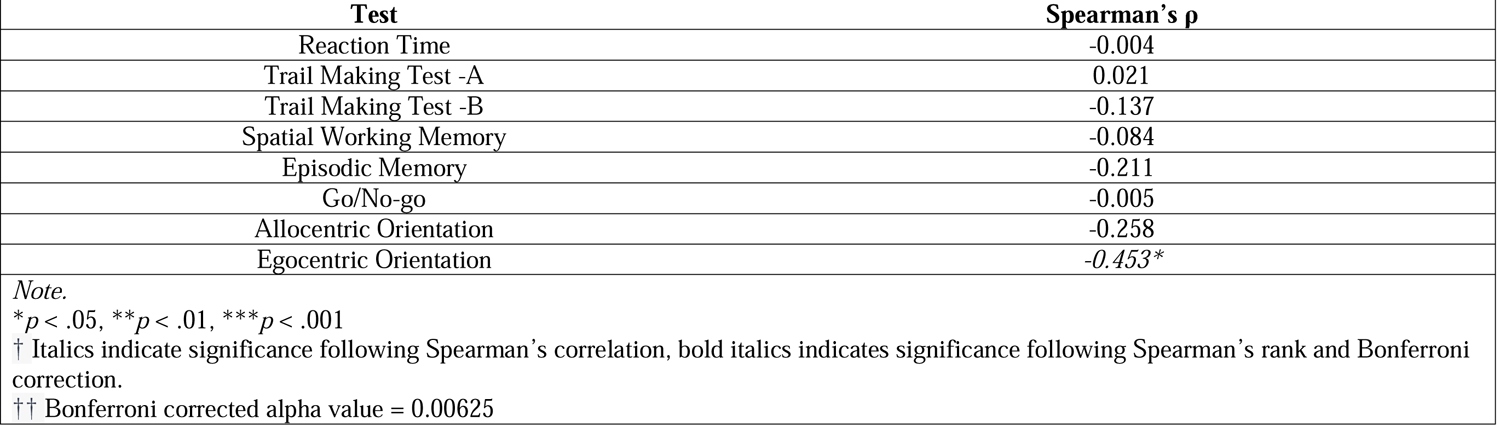
Correlation analysis between online cognitive testing and the CCI.

Correlation analysis was then conducted to establish whether global cognitive performance from the online cognitive battery validated against the MoCA. A Pearson’s correlation found that global cognition performance showed a moderate negative correlation with MoCA performance, r(24) = .603, *p* = .001 (see Figure 1).

**Figure 1.**
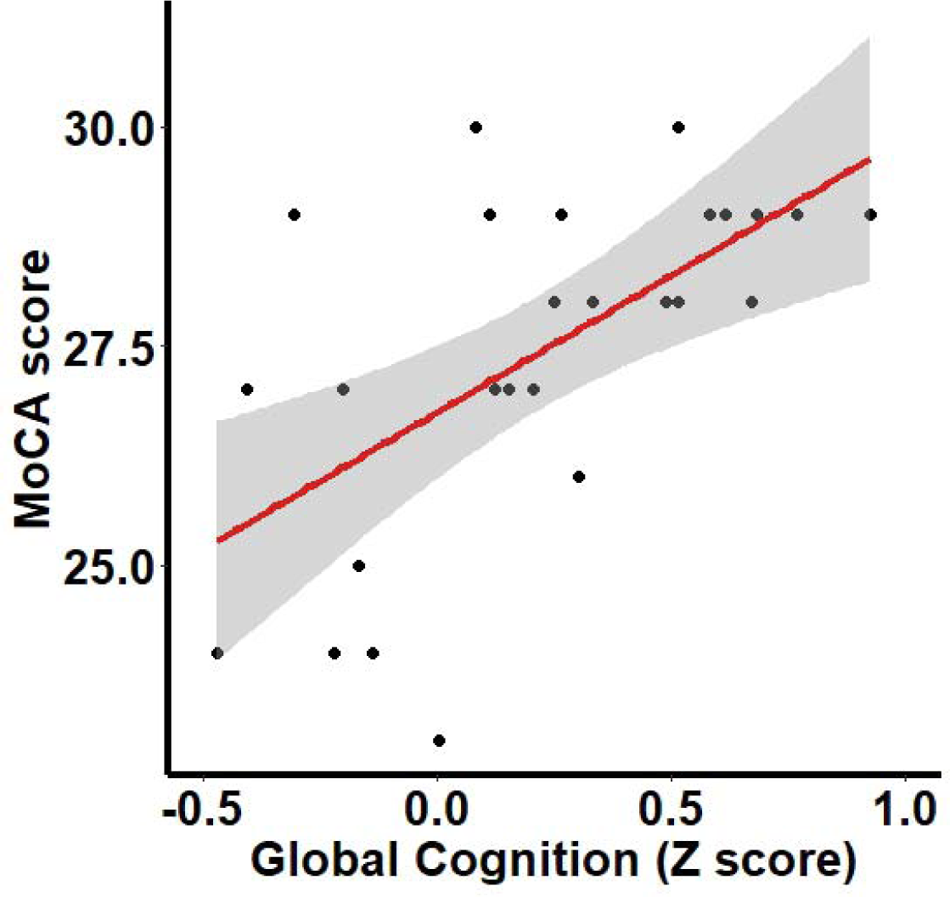
Regression plot showing the relationship between MoCA score and global cognitive performance in the NeurOn battery.

### Influence of factors on neuropsychological testing

To explore how demographic factors influenced performance in the online cognitive battery, a multiple linear regression analysis was conducted using the variables of traditional test scores, age, sex, and education. Traditional test scores significantly predicted performance in Trail Making Test - B performance (β = 0.49, *p* = 0.002, CI[0.20, 0.78]) and Global Cognition (β = 0.589, *p* < 0.001, CI[0.33, 0.84]). Older age was associated with worse performance in Trail Making Test - A (β = −0.12, *p* = 0.030, CI[−0.23, −0.01]) and allocentric orientation (β = 0.10, *p* = .007, CI[−0.16, −0.03]). Sex was positively associated with episodic memory, with being female predicting better episodic memory performance (β = 7.759, *p* = .02). More years in education was associated only with allocentric orientation (β = 0.09, *p* < .05., CI[0.00, 0.17]) (see Table 4).

**Table 4.** Multiple linear regression analysis of factors associated with online cognitive tests.

|  |  |  |  |  |  |
| --- | --- | --- | --- | --- | --- |
| **Test** | **Traditional test** | **Age** | **Sex** | **Education** | **Model R<sup>2</sup>** |
| Reaction Time | - | -0.01 | -0.65 | 0.01 | .12 |
| Trail-Making Test A | 0.20 | **-0.12\*** | 0.19 | 0.02 | .25 |
| Trail-Making Test B | **0.49\*\*** | -0.04 | -0.11 | 0.07 | .51 |
| Spatial Working Memory | 0.22 | -0.07 | -0.25 | -0.02 | .20 |
| Episodic Memory | 0.05 | 0.01 | **0.12\*** | 0.01 | .24 |
| Go/No-Go | - | 0.03 | 0.58 | 0.06 | .13 |
| Allocentric Orientation | - | **-0.10\*\*** | -0.26 | **0.09\*** | .33 |
| Egocentric Orientation | - | -0.01 | 0.38 | 0.06 | .10 |
| Global cognition | **0.58\*\*\*** | -0.01 | -0.08 | -0.02 | .76 |
| *Note.*<br>*$p < .05$, **$p < .01$, ***$p < .001$<br>† Displaying unstandardised beta coefficients.<br>†† Cognitive tests were standardised and reverse scored for analysis so positive score reflects better performance. |  |  |  |  |  |

## Discussion

With the rising aging population, there is an urgent need to establish screening tools for early identification of cognitive decline in aging. This study assessed the feasibility, reliability, and validity of a novel online cognitive testing battery in an older adult population to establish its applicability in acquiring cognitive performance data in a healthy older adult population within unsupervised, remote settings. Importantly, we demonstrate that global performance in the cognitive battery validates against the MoCA – one of the most popular tests for screening for mild cognitive impairment (MCI). We also demonstrate that egocentric orientation performance was the only cognitive domain associated with ratings on the CCI. As predicted, we establish test-retest reliability of the battery as all repeated tests showed moderate test-retest reliability and no practice effects were present after a one-week washout period between testing sessions. Lastly, we explore factors that influence performance in online cognitive assessments and found that age positively correlates with processing speed and allocentric orientation performance.

Due to individual differences in cognitive trajectories in ageing, composite measures assessing a range of cognitive domains have been suggested as the most appropriate approach to screen for and track cognitive impairment over time (32). Within the present study, we demonstrate that global cognitive performance in the online cognitive battery shows a strong correlation with global cognition measured by the MoCA. To date, very few studies have validated online cognitive assessments in older adults (33), with few showing that online cognitive assessments provide similar diagnostic accuracy to the MoCA (34). Our results show that the NeurOn battery provides a promising instrument for measuring cognitive performance remotely at a similar accuracy with clinical testing appointments.

Many traditional cognitive assessments, such as the MoCA and MMSE, are limited by practice effects which may compromise the ability to interpret whether cognitive change is due to task experience rather than aging effects (35,36). Practice effects are more likely to occur within shorter testing intervals and are prominent across one week re-testing intervals (37,38). In the present study, despite a short retesting period of one week, no statistically significant improvement was found over any of the repeated tasks. This is potentially due to the pseudorandomisation of task material within the NeurOn battery, which enables the task to regenerate so that material cannot be learned by participants. Although a one-week retesting period is not typically used for clinical relevance for neuropsychological testing (38), it is potentially useful for assessing cognitive changes after short-term intervention studies (39). Reduced practice effects also enable for identification of subtle changes in cognitive trajectories longitudinally, where currently they are rarely conducted in routine clinical appointments due to being resource intensive.

Contrary to our hypotheses, we found that only the Trail Making Test - A and global cognitive performance demonstrated concurrent validity to the paper-based tasks, respectively. Previous research has found that concurrent validity of online cognitive testing is typically low (median 0.49) (17), and therefore correspondence between online cognitive tests and paper-based tasks is often at best moderate. It is possible that digitalising some traditional paper-based tasks influences test performance and therefore comparing online cognitive test performance to normative data in non-computerised tasks may not be valid in assessing for cognitive impairment. However, due to the greater precision, standardisation, and objectivity in data measurement for online cognitive testing, computerised cognitive tasks can be used to develop new normative data cut-offs that can assess more sensitively for cognitive changes. Furthermore, online testing opens the possibility of testing a significantly larger and wider population demographic who may not be able to receive clinical assessments. By establishing large normative datasets, it is possible to establish how cognitive changes over time differ across specific subpopulations, which enable for more accurate diagnostic markers (40). Age-related variability in cognitive performance increases rapidly after age 60 (41), and therefore it is necessary to establish sociodemographic factors that may influence interpretation of cognitive trajectories.

In the present study, we found that egocentric orientation was the only cognitive test found to correlate with CCI score, which is commonly used to identify subjective cognitive decline (SCD) (42). Previous research has established that individuals with SCD typically show spatial orientation deficits (43,44), although little is known about how this relates to performance across other cognitive tasks. The present findings indicate that egocentric orientation deficits may be a key signature for SCD, supporting growing findings that spatial orientation performance as a marker for early cognitive impairment (21). SCD typically manifests prior to preclinical dementia (45), yet there is large heterogeneity in the outcomes of SCD, with many individuals experiencing SCD without objective cognitive impairments (46). As the only test correlating with worse subjective cognition was egocentric orientation, future research may look to establish whether individuals with SCD who exhibit worse egocentric orientation abilities may be more at-risk for future cognitive impairment.

Overall, the novel cognitive battery demonstrates the usability and feasibility in measuring cognitive performance remotely, as all participants were able to complete the assessment unsupervised at home using a variety of devices. Our battery has also previously demonstrated feasibility and internal consistency in collecting large longitudinal normative cognitive data across regions (unpublished data). Many cognitive assessments, such as the MoCA, are limited in their generalisation across different cultures as they require the use of language and cultural understanding (47). A strength in the NeurOn battery is that tasks are visual and do not require language, which enables for greater cross-cultural generalisation in cognitive performance and meets the ambitions of in improving global dementia screening (48). Although online cognitive testing has several advantages over in-person clinical assessments, diagnosing cognitive impairments, such as MCI, requires a functional and clinical evaluation (49) and therefore should not take place outside of a clinical setting. Presently, online cognitive testing may offer a potential pre-screening tool for extensive clinical assessments, such as neuroimaging and biomarker testing. Furthermore, online cognitive testing can advance research by increasing the scale of epidemiological studies (50) as well as screening participants for participation in clinical trials (51).

Although our results are promising, this study has some limitations. We did not account for computer skill, which has previously been found to relate to better cognitive task performance (18,52). Secondly, our sample size of 32 was relatively low and therefore more work is required to comprehensively understand how sociodemographic factors influence neuropsychological tests within the NeurOn battery. Our sample only consists of healthy individuals, and therefore there is currently little understanding as to the feasibility of the NeurOn battery within patient population groups. Research is currently ongoing to examine how NeurOn test performance differs across healthy ageing, preclinical dementia, and early dementia. Lastly, unsupervised cognitive testing naturally has drawbacks in that there is a lack of standardisation in home testing, and therefore it is possible that participant performance at home may be influenced by confounding factors i.e., distraction. However, participants were provided with clear instructions to mitigate this.

In conclusion, the NeurOn cognitive assessment battery demonstrates a promising instrument for assessing cognitive performance within healthy older adult populations. In the present study, the NeurOn battery compared well with MoCA performance; showed negligible practice effects; and was easily administered in unsupervised remote testing environments. Future research in online cognitive assessments should look to establish appropriate testing timepoints for sensitively measuring longitudinal changes in cognitive functioning.

## Data Availability

All data produced in the present study are available upon reasonable request to the authors following publication.

## Acknowledgments

This study is supported by the National Institute for Health and Care Research (NIHR) Applied Research Collaboration East of England (NIHR ARC EoE) at Cambridge and Peterborough NHS Foundation Trust. The views expressed are those of the author(s) and not necessarily those of the NIHR or the Department of Health and Social Care. SM’s studentship is jointly funded by the Faculty of Medicine & Health Sciences, UEA, and the Earle and Stuart Charitable Trust.

